# Phenotypic Severity of *SCN5A* Related Bradycardia is Independent of Dominant-Negative and Coupled Gating Effects

**DOI:** 10.1101/2025.08.06.25332682

**Authors:** Ayami Tano, Koichi Kato, Kohei Yamauchi, Hideyuki Jinzai, Futoshi Toyoda, Yuichi Baba, Toru Kubo, Seiko Ohno, Takeru Makiyama, Yoshihisa Nakagawa, Minoru Horie

## Abstract

**Background:** Pathogenic *SCN5A* variants are associated with inherited arrhythmias such as long QT syndrome, Brugada syndrome, and sick sinus syndrome (SSS). While Na_v_1.5, an α-subunit of the cardiac sodium channel encoded by *SCN5A*, has been considered to function as a monomer, recent studies reveal that a reduction of sodium current in wild-type (WT) Na_v_1.5 can be caused by dimerization with loss-of-function (LOF) mutated Na_v_1.5 through dominant-negative (DN) effects. However, the clinical significance of the DN effect remains unclear.

**Method:** We genetically screened a family who presented with SSS and sudden cardiac death (SCD). Whole-cell patch-clamp study using HEK293 cells co-expressing WT- and variant SCN5A was performed. Channel dimerization was assessed by co-immunoprecipitation and proximity ligation assays. Also, the effects of difopein, a high affinity inhibitor of Na_v_1.5 interaction via 14-3-3 proteins, were evaluated.

**Results:** The proband carried compound heterozygous variants p.T1396P and p.G833R. The whole-cell mode patch-clamp techniques demonstrated that the p.T1396P showed a DN effect on the peak sodium currents (37% decrease in I_Na_) and altered gating properties (+5.6 mV shift in steady-state inactivation) when expressed with WT *SCN5A*. These effects were abolished by difopein. p.G833R showed no DN or coupled gating effect but still formed dimers. The proband developed earlier and more severe bradycardia than her parent, who only carries p.T1396P, suggesting that loss of coupled gating effect contributed to the severe phenotype.

**Conclusion:** Our findings suggest that coupled gating may be physiologically important for normal Na_v_1.5 function, and that its loss can exacerbate disease severity.

## Introduction

The voltage-gated sodium channels are expressed in various excitable cells and are essential for the generation and propagation of action potentials^1^. Among them, the human Na_v_1.5, encoded by the *SCN5A* gene, plays a crucial role in cardiac excitability. Pathogenic variants in *SCN5A* are associated with a spectrum of inherited arrhythmic diseases, including long QT syndrome, Brugada syndrome (BrS), and sick sinus syndrome (SSS)^2,3,4^.

Unlike potassium channels that form functional oligomers, the α-subunit of Na_v_1.5 has long been considered to form an ion pore as a monomer. However, recent studies show that co-expression of wild-type (WT) and loss-of-function (LOF) type of the *SCN5A* variants can reduce peak sodium currents. This phenomenon, known as a dominant-negative (DN) effect, suggests that LOF variants can interfere with WT channel function. Adding to that, the coupled-gating effect has been also suggested through the demonstration that voltage-dependance of WT channel was affected by LOF variants. The DN and coupled-gating effect are reported to be mediated through channel dimerization, facilitated by 14-3-3 protein interactions^5,6,7,8^. Despite these findings, the clinical relevance of the DN effect has yet to be uncovered. A previous study reported a higher penetrance in affected families carrying DN variants, but no clear association with phenotype severity was observed^8^. Thus, we aim to find DN *SCN5A* variants in various genetic arrhythmias. In this study, we found a family who showed SSS and SCD harboring two *SCN5A* variants: p.T1396P and p.G833R. The patch-clamp study revealed that p.T1396P, but not p.G833R, showed a DN and coupled-gating effect. We further investigated the effects of these variants on activation and inactivation kinetics. In addition, we also assessed the association between disease phenotype severity and the presence of these effects.

## Method

This study was approved by the institutional review board of Shiga University of Medical Science (G2011-128). All patients provided written informed consent before genetic investigations in accordance with the Declaration of Helsinki.

### Genetic analysis

Genomic DNA was extracted from the patients’ blood lymphocytes using standard protocols. Coding exons of known pathogenic genes of inherited arrhythmias^9^ were screened using targeted short-read sequencing (targeted SRS) with the HaloPlex HS custom panel (Agilent Technologies, Santa Clara, CA, USA) and MiSeq (Illumina, San Diego, CA, USA). Data analyses were performed using SureCall software (Agilent Technologies, Santa Clara, CA, USA). Detected variants were confirmed by Sanger sequencing. Genetic variants were classified based on the American College of Medical Genetics and Genomics (ACMG) guidelines^10^.

### Genetic engineering

For the electrophysiological study and cell-surface biotinylation assay, the full-length cDNA of the *SCN5A* encoding the human Na_v_1.5 channel was sub-cloned into a pcDNA3.1(+) vector (Thermo Scientific, Waltham, Massachusetts, USA). Mutants were generated by site-directed mutagenesis using the QuickChange-II-XL kit (Stratagene, La Jolla, CA, USA) according to the manufacturer’s instructions. For the co-immunoprecipitation assay and the proximal ligation assay, HA tag sequence and FLAG sequence were introduced in the pcDNA3.1-*SCN5A* plasmid sequence.

### Cell preparation

Human embryonic kidney 293 (HEK293) cells were cultured and passaged using a complete medium consisting of DMEM, 4.5 g/L glucose, 5% fetal bovine serum (FBS), and 1% Penicillin/Streptomycin solution (Nacalai Tesque Inc., Kyoto, Japan) in a humidified incubator at 37°C with 5% CO_2_. Transfection was performed using JetPEI (Polyplus-transfection, Illkirch, France) according to the manufacturer’s instructions.

### Electrophysiological assay

HEK 293 cells seeded in 35mm dishes were transfected with various combinations of WT- and mutant-*SCN5A* plasmids. Forty-eight hours after transfection, I_Na_ was measured *in vitro* using the EPC-8 patch-clamp amplifier in a whole-cell patch-clamp mode (HEKA Elektronik, Lambrecht, Germany). The cells were perfused in an adjusted solution (composition: 80.0 mM NaCl, 2.0 mM CaCl_2_, 10.0 mM HEPES, 2.5 mM MgCl_2_(6H_2_O), 50.0 mM CsCl, adjusted in pH 7.4 with CsOH) kept at 37°C in a recording chamber on the stage of a microscope. A micropapillary (WPI, FL, USA) was pulled to prepare patch-electrodes using a microelectrode puller (P-97, Sutter Instrument Co., Novato, CA, USA). The patch-electrode kept a resistance of 1.0-1.5 MΩ when it was filled with an electrode solution (composition: 5.0 mM NaCl, 2.0 mM MgCl_2_ (6H_2_O), 1.0 mM CaCl_2_, 15.0 mM EGTA, 10.0 mM HEPES, 1.0 mM MgATP,

130.0 mM CsCl, adjusted in pH 7.2 by CsOH). The recording protocols (activation, inactivation, and recovery) were all programmed on the computer software Patchmaster (HEKA Elektronik, Lambrecht, Germany). The membrane capacitance was measured for each cell and used for compensation of the each recorded I_Na_ value. The Na_v_1.5 conductance (*G*) was calculated using the equation [*G = I*o*/*(*V – E*_rev_)] (*I*o: peak I_Na_ amplitude, *V:* test potential, *E*rev: reversal potential). Voltage-activation relationships of *I*_Na_ were evaluated by fitting the relative conductance-Vm relationships to a Boltzmann equation: G/Gmax = 1/[1 + exp(-[*V_m_* –*V_h_*]/*k*)], where G/Gmax is the relative conductance, V1/2 is the voltage at half-maximal activation, Vm the test potential, and *k* the slope factor. The time dependent decay of *I*_Na_ after depolarization was determined by single exponential fits of current traces.

### Cell surface biotinylation

HEK 293 cells seeded in 60 mm dishes were transfected with WT- and mutant-*SCN5A* plasmids. Forty-eight hours after the transfection, cells were washed twice with ice-cold PBS and incubated at 4°C with PBS containing 1.5 mg/mL of EZ Link Sulfo-NHS-SS-Biotin (Catalog #21331, Thermo Fisher Scientific, Waltham, MA, USA). After 60 min, the plates were washed twice and incubated for 10 min at 4°C with PBS and 100 mM glycine to quench unlinked biotin, then washed twice with PBS. Cells were then lysed with lysis buffer (10 mM HEPES, 150 mM NaCl, 1 mM EDTA, 1% Nonidet P-40, proteinase inhibitor cocktail) and collected in 1.5 ml tubes. After centrifugation at 14,000 g for 30 min at 4°C, part of the supernatants was kept for total protein evaluation. Residual supernatants were incubated with immobilized neutravidin beads (Pierce, USA) overnight at 4°C and pelleted by centrifugation at 14,000 g for 1 min at 4°C. After three washes with lysis buffer, the biotinylated proteins were eluted with 10ul of Low pH solution (0.1 M HCl 1 ml, 0.2 M DTT 2 ml) at room temperature for 30 minutes. After adding 10 µl of 4x NuPAGE LDS Sample buffer (Thermo Fisher Scientific) and 5 µl of Tris solution (0.5 M Tris, 0.4% SDS, pH adjusted to 6.8), biotinylated proteins were analyzed using Western blot.

### Co-immunoprecipitation assay (CO-IP assay)

HEK 293 cells cultured in 60 mm dishes were transfected with WT and mutant-*SCN5A* plasmids (3µg). Forty-eight hours after transfection, cells were washed, lysed, and collected by the same method above mentioned. After centrifugation at 14,000 g for 30 min at 4°C, part of the supernatants was kept for total protein evaluation. Residual supernatants were incubated with anti-HA antibody (#3724, Cell signaling, Danvers, MA, USA) in a dilution ratio of 1: 200 (Ab: lysate) on a rotator overnight at 4°C. Immunoprecipitation was performed using DynabeadsTM protein G kit (Catalog # DB10007, Thermo Fisher Scientific, Waltham, MA, USA), according to the manufacturer’s protocol. The precipitated protein was analyzed by using western blotting.

### Western blotting analysis

The eluted samples through the biotinylation and CO-IP above were mixed with sample loading buffer and processed using gel-electrophoresis at 100V for 3 hours. The proteins in the gel were transferred to PVDF membranes using the Trans-blot Turbo blotting system (BIO-RAD, Berkeley, CA, USA) at 1.0 V (25 mA) for 30 minutes. After one hour of blocking with phosphate buffered saline with Tween 20 solution (PBS-T) including 5% skim milk, the membranes were incubated in diluted first-antibodies (Anti FLAG: F3165, Sigma, USA; Anti GAPDH: ab9485, abcam, Cambridge, UK) overnight at 4°C. The membranes were washed with PBS-T and incubated with second-antibodies for one hour at room temperature. The membranes soaked in a chemiluminescent solution (Chemilumi-one, Nacarai, Kyoto, Japan) were captured using a chemiluminescence imager, ImageQuantTM LAS-4000 (Cytiva, Marlborough, MA, USA).

### *in situ* proximal ligation assay

To visualize the localization and the proximity of Na_v_1.5 channels, we performed *in situ* proximal ligation assay using Duolink PLA kit (Sigma-Aldrich, St. Louis, MO, USA). HEK 293 cells seeded on round cover glass slides were transfected with HA- or FLAG-tagged WT and mutant-*SCN5A* plasmids in various combinations. Forty-eight hours after transfection, cells were washed twice with PBS. The cells were covered by 1 ml of 4% paraformaldehyde (PFA) and incubated on an agitator for 10min in room temperature. After washing by PBS three times to remove PFA, 0.1% Triton-X100 (1 ml) was added to permeabilize the cells. After two minutes of incubation on an agitator, cells were washed by PBS three times. The slides were covered by 40 µl of Duolink blocking solution and incubated for 1 h in 37[. After tapping off the blocking solution, first antibodies (Anti HA: #3724, Cell signaling; Anti FLAG: F3165, Sigma) diluted in Duolink Antibody Diluent were added to the slides. Cells were incubated in a humid chamber at 37 [overnight. Then, PLA probe reaction, ligation, and amplification steps were performed following the manufacturer’s protocol. The slides were observed by a confocal laser scanning microscope TCS SP8 X (Leica, Wetzlar, Hessen, German). The PLA signals indicate the colocalization (<40nm) of two proteins, each labeled with a distinct tag and detected by specific antibodies. The area of PLA signals per cell were evaluated and analyzed.

### Statistical Analysis

Data were shown as mean ± SEM or ± SD when specified. Statistical significance was evaluated by one-way ANOVA and Dunnett’s multiple comparison test. *p* < 0.05 was considered significant.

## Results

### Clinical characteristics

Figure 1A shows the pedigree of our patients. The proband is a female in her 30s who experienced a sudden loss of consciousness late at night. She presented with recurrent episodes of ventricular tachycardia (VT) and severe sinus bradycardia (Figure 1B). In the acute phase, her echocardiography revealed an aneurysmal change in the left ventricular apex, which was diagnosed as Takotsubo cardiomyopathy. Although the echocardiographic abnormality was completely resolved over time, the patient’s sinus bradycardia persisted into the chronic phase, and an implantable cardioverter defibrillator (ICD) was implanted due to the initial VT. Since the device implantation, she has remained free from VT recurrence. One of her parents also had a history of sinus bradycardia in their 40s after taking a class I antiarrhythmic agent due to paroxysmal atrial fibrillations. Shortly after administration, the individual was unconscious due to sinus bradycardia, and temporary cardiac pacing was performed. Their sinus rhythm subsequently recovered without a permanent pacing (Figure 1B). The proband’s sibling was found to have bradycardia and left ventricular enlargement at school based routine ECG screening in childhood. This patient underwent a pacemaker implantation by age 10. In their late teens, the individual died suddenly, possibly due to tachycardia, though the electrogram data from the pacemaker was unavailable. The proband’s another sibling and the other parent have no history of arrhythmic events and their ECGs are normal.

**Figure 1.**
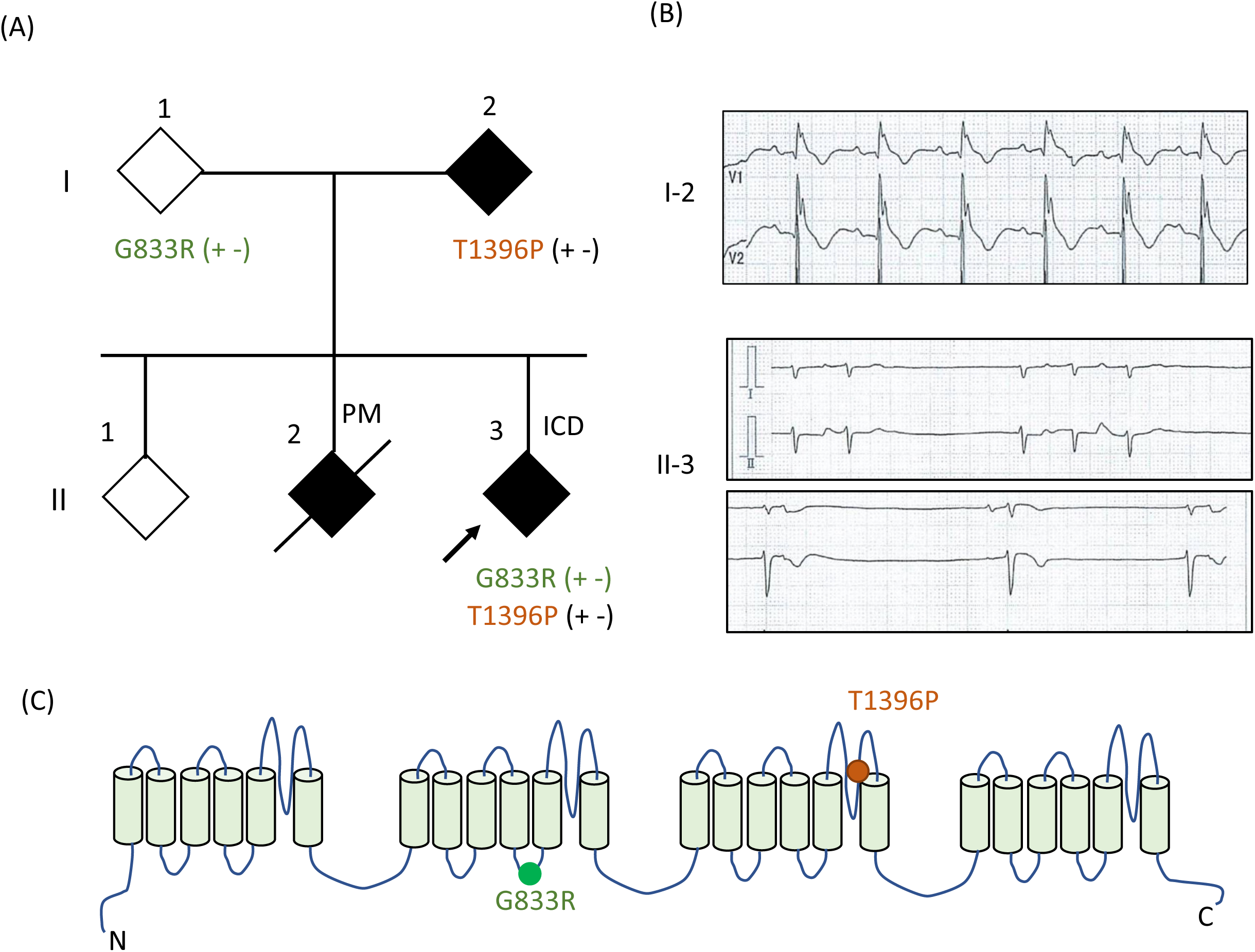
A family pedigree and the SCN5A variants A) Pedigree of an affected family harboring two *SCN5A* variants. Filled symbols indicate individuals with bradyarrhythmia. B) ECG recordings of individuals I-2 and II-3. C) Topology of Nav1.5 indicating the locations of the two identified variants.

### Identification of *SN5A* variants

We identified two *SCN5A* variants, c.2497 G>A (p.G833R) and c.4186 A>C (p.T1396P), in the proband in a compound heterozygous fashion (Figure 1A**, C**). The minimum allele frequency (MAF) of p.G833R is 0.029% in East Asian, and p.T1396P has not been found in general populations according to the gnomAD v4.1.0 (https://gnomad.broadinstitute.org/gene/ENSG00000183873?dataset=gnomad_r4). One of her parents carried only the heterozygous p.T1396P, and the other parent carried the heterozygous p.G833R. The genomic sample from the proband’s deceased sibling was unavailable. According to the American College of Medical Genetics and Genomics (ACMG) guidelines^8^, both variants were classified as variants of unknown significance (VUS).

### Electrophysiological characteristics of the variant channels

Patch-clamp analysis was performed in HEK 293 cells transfected with various combinations of WT- and mutant-*SCN5A* plasmid (Figure 2**, 3** and **Table**). The transfection conditions are shown in Figure 2A. The amounts of plasmids were 0.5 µg for each type (i.e., WT and variants), and the total amount of plasmid was adjusted to 1.5 μg with pEGFP-C3 plasmid. When only *SCN5A* p.T1396P plasmids (0.5 µg) were transfected, the cell did not generate I_Na_, indicating that p.T1396P is a loss-of-function (LOF) variant (Figure 2B**, filled purple triangle**). In the cells co-transfected with WT and p.T1396P plasmids (0.5 µg each), the peak I_Na_ decreased by 37% (Figure 2B**, pink triangle**) compared to the cells transfected with WT plasmid (0.5 µg). This result suggested that the p.T1396P variant showed a dominant negative effect.

**Figure 2.**
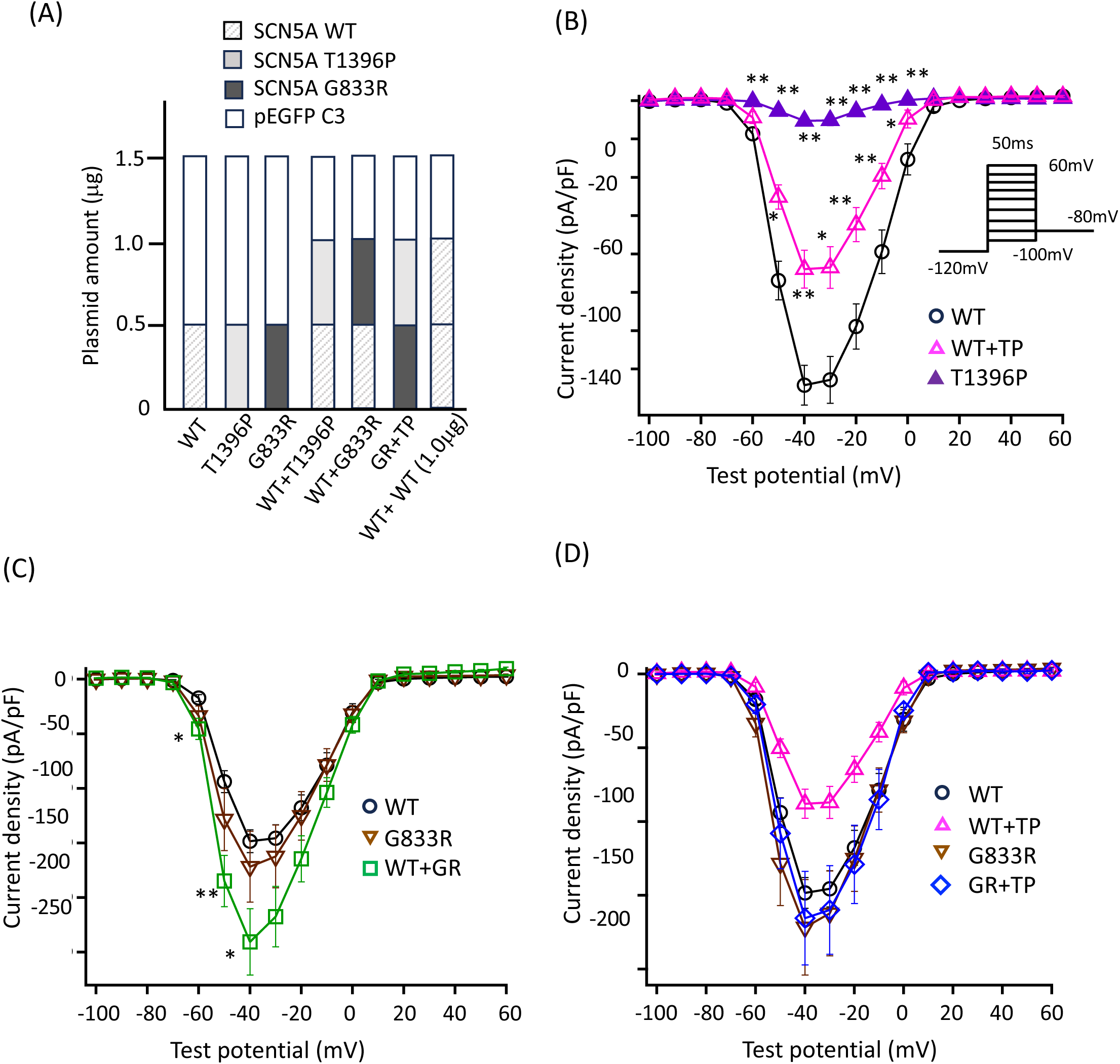
Electrophysiological analysis of SCN5A variants A) Transfection conditions. In all groups, the total plasmid amount was adjusted to 1.5[µg. The meaning of each shading pattern in the bar graph is shown in the legend inset. B–D) Current–voltage (I–V) relationships for cells expressing various plasmid combinations. The test pulse protocol is shown in the inset. *\* p* < 0.05; ** *p* < 0.01. WT (wild-type) by one-way ANOVA followed by Tukey’s post hoc test vs WT. TP (T1396P); GR (G833R).

In contrast, the I_Na_ in the cells transfected with p.G833R (0.5 µg) was comparable to that of WT (Figure 2C**, brown inverted triangle**). When p.G833R (0.5 µg) was co-transfected with WT (0.5 µg), the peak I_Na_ was significantly larger than cells transfected with WT (0.5 µg) alone (Figure 2C**, green square**) and comparable in the cell transfected with WT (1.0 µg) (**supplemental figure A, Table**). Interestingly, when p.G8333R (0.5 µg) was co-transfected with p.T1396P (0.5 µg), the peak I_Na_ did not decrease (Figure 2D, blue diamond), indicating that p.T1396P did not exhibit dominant-negative effect on the p.G833R channels.

We further investigated the effects of p.T1396P on various characteristics of the WT channel. When p.T1396P plasmids (0.5 µg) were co-transfected with WT plasmids (0.5 µg), the fast component of current decay was prolonged, and the voltage-dependence of steady-state inactivation was shifted rightward by 5.6mV (Figure 3A**, B, Table**), while the voltage-dependence of activation was not affected (Figure 3C).

**Figure 3.**
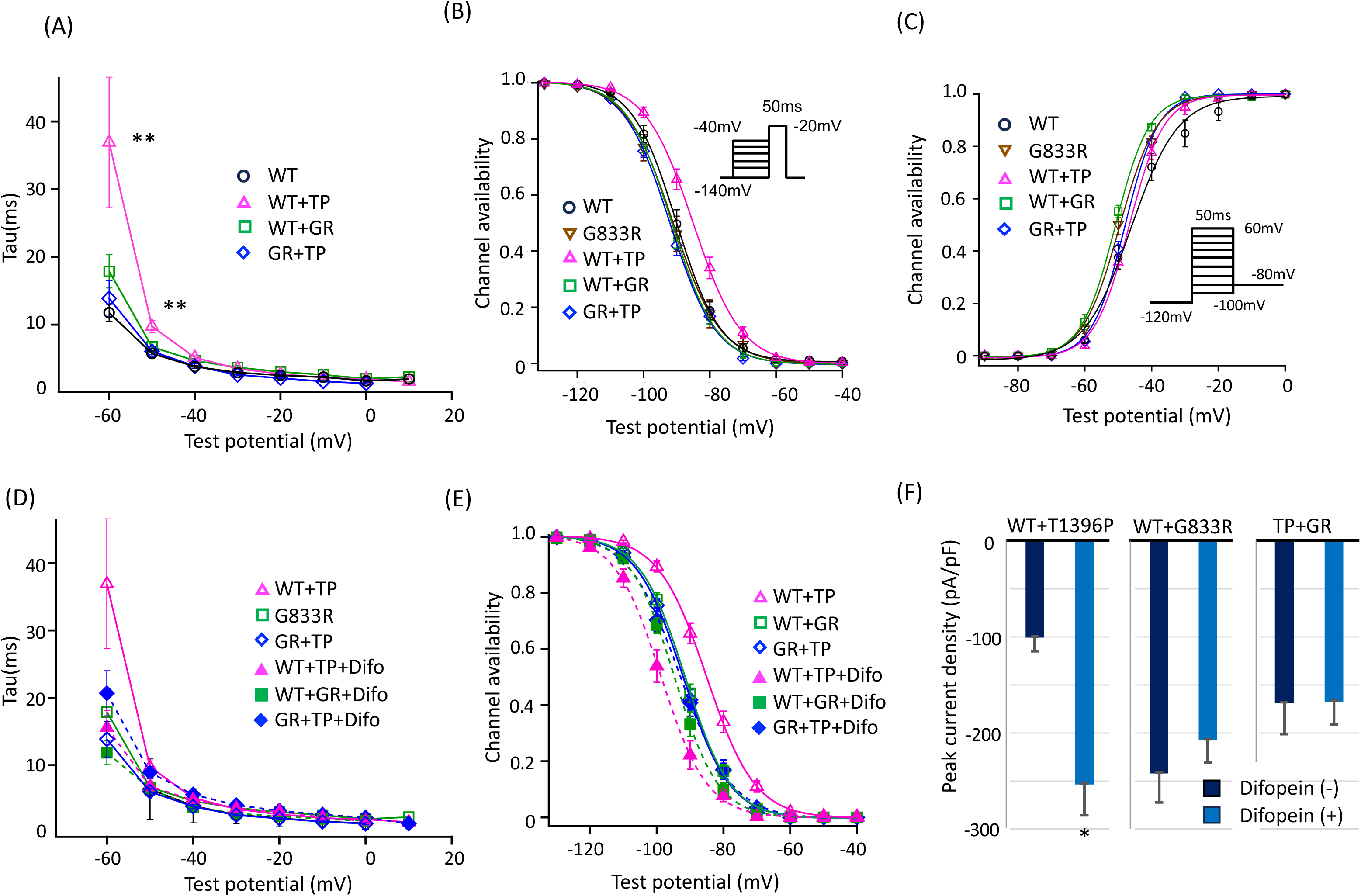
Electrophysiological properties of SCN5A variants A) Time constant of current decay for various test potentials. B, C) Steady-state inactivation and activation curves. The test pulse protocol for inactivation is shown in inset. D) Time constant of current decay in cells co-expressing difopein. E) Steady-state inactivation curve in cells co-expressing difopein. F) Bar graphs representing peak I_Na_ density at -40mV with or without co-expressing difopein. *\* p* < 0.05; ** *p* < 0.01 by one-way ANOVA followed by Tukey’s post hoc test vs WT. WT (wild-type); TP (T1396P); GR (G833R).

Despite p.T1396P channel being non-functional, it affected WT channels, indicating these two channels can function by interacting each other (coupled gating).

### Difopein prevented the interactions between WT and variant Na_v_1.5

Previous studies reported that difopein inhibits 14-3-3 proteins that play a role in the dimerization of Na_v_1.5 channels^6,11^. To investigate this further, we co-expressed this peptide with various combination of WT and variant SCN5As . Co-expression of difopein with WT and p.T1396P channels reversed the prolonged current decay and rightward-shift of the steady-state inactivation curve. In contrast, difopein did not affect these parameters in cells transfected with p.G833R (Figure 3D**, E**). In addition, the steady-state inactivation for cells expressing only WT was also shifted leftward by co-expression of difopein (Supplemental figure B).

Furthermore, the peak I_Na_ in the cells transfected with WT and p.T1396P plasmids increased by 1.5 fold (Figure 3F) with difopein co-expression. In contrast, difopein did not affect the I_Na_ in the cells expressing WT and p.G833R, or transfected with p.G833R and p.T1396P. In cases where a dominant negative effect was not observed, co-expression of difopein did not increase current densities, suggesting the increase of the peak current density is via cancellation of dominant-negative effect by T1396P.

### Comparison of cell-surface expression of WT and each variant Na_v_1.5

To assess the cell-surface expression of each mutated Nav1.5, we performed a cell-surface biotinylation assay (Figure 4A**, B**). The quantitative analysis of cell-surface expression exhibited no significant difference among WT and two variant channels, suggesting that both p.G833R and p.T1396P variants do not affect translation and membrane trafficking of Na_v_1.5.

**Figure 4.**
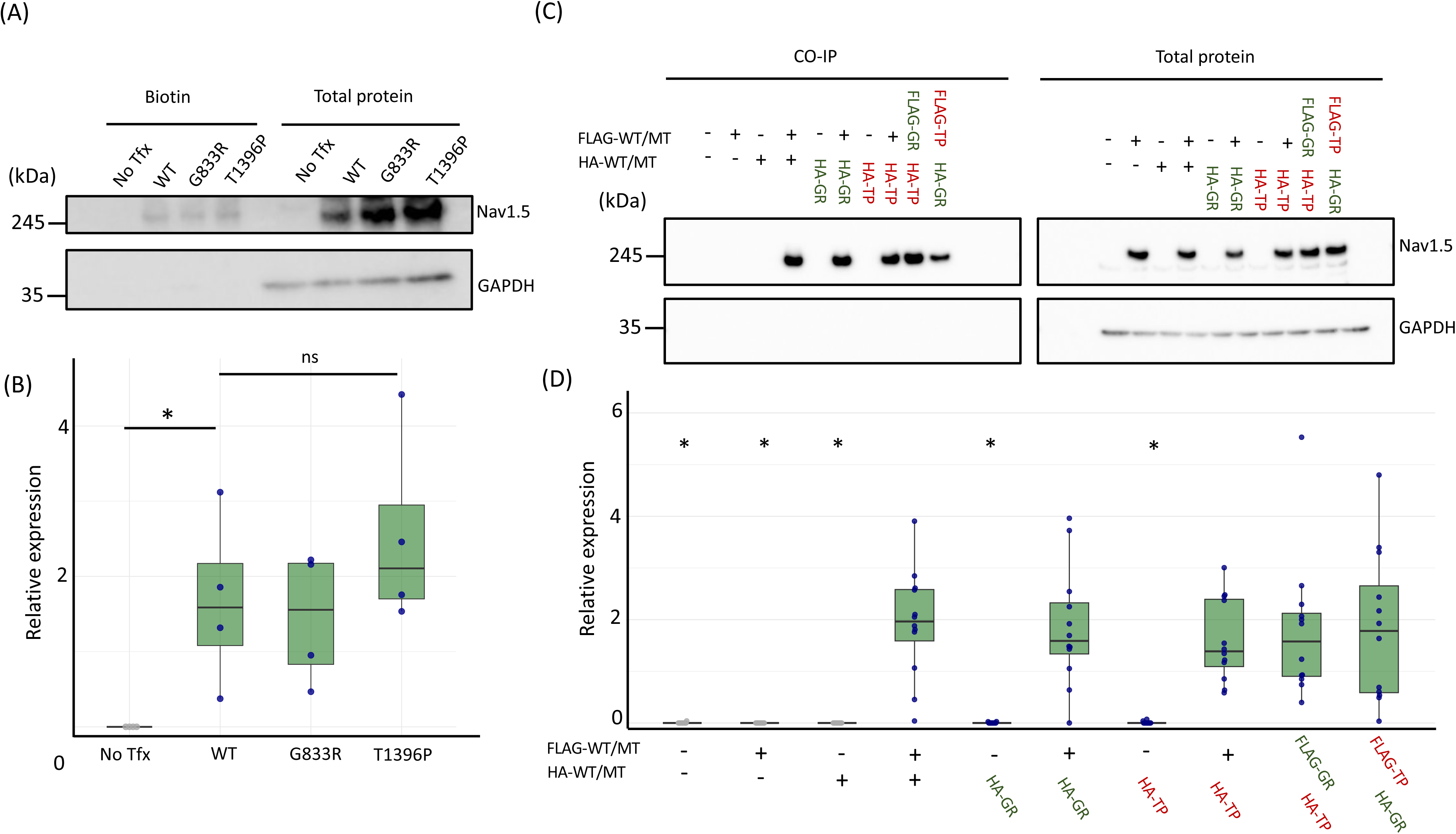
Biotinylation and co-immunoprecipitation assays. A) Representative immunoblot images of biotinylated surface protein fractions and total protein fractions from cells transfected with wild-type or mutant SCN5A plasmids. B) Beeswarm plots showing individual data points from the biotinylation assay (n = 4). Box plots are overlaid to indicate the median, interquartile range (box), and data distribution (whiskers). C) Representative immunoblot images from the co-immunoprecipitation (CO-IP) assay. Shown are the Co-IP fractions and corresponding total protein fractions from cells transfected with various plasmid combinations. **D)** Beeswarm plots with overlaid box plots representing individual data points from the Co-IP assay (n > 7). “No Tfx” indicates non-transfected cells. FLAG-WT/MT (FLAG-tagged wild-type/mutant); HA-WT/MT (HA-tagged wild-type/mutant); FLAG-GR/TP (FLAG-tagged G833R/T1396P); HA-GR/TP (HA-tagged G833R/T1396P). \**p* < 0.05; \*\**p* < 0.01 by one-way ANOVA followed by Tukey’s post hoc test vs WT.

### WT and variant Na_v_1.5 assembled into oligomers, irrespective of their electrophysiological characteristics

Next, to evaluate the oligomerization patterns of WT and variant channels, we performed co-immunoprecipitation (co-IP) assays using HA-tagged and FLAG-tagged WT and variant channel plasmids. Figure 4 shows that the HA-tagged Na_v_1.5 was co-precipitated by the FLAG-tagged Na_v_1.5, regardless of the electrophysiological effects. This result indicates that Na_v_1.5 form oligomers.

### *In situ* proximity ligation assay exhibited close localization of WT and mutated Na_v_1.5

To further assess the assembly of Na_v_1.5 in more physiological conditions, we performed an *in situ* proximity ligation assay (Figure 5) using HA- and FLAG-tagged constructs. Through the microscopic evaluation, we observed PLA signals in the cells transfected with all pairwise combinations of *SCN5A* plasmids (Figure 5A) irrespective of the electrophysiological characteristics, suggesting the proximal expression of each tagged channels in living cells. This result supports the co-IP results showing that Na_v_1.5 channels form oligomers.

**Figure 5.**
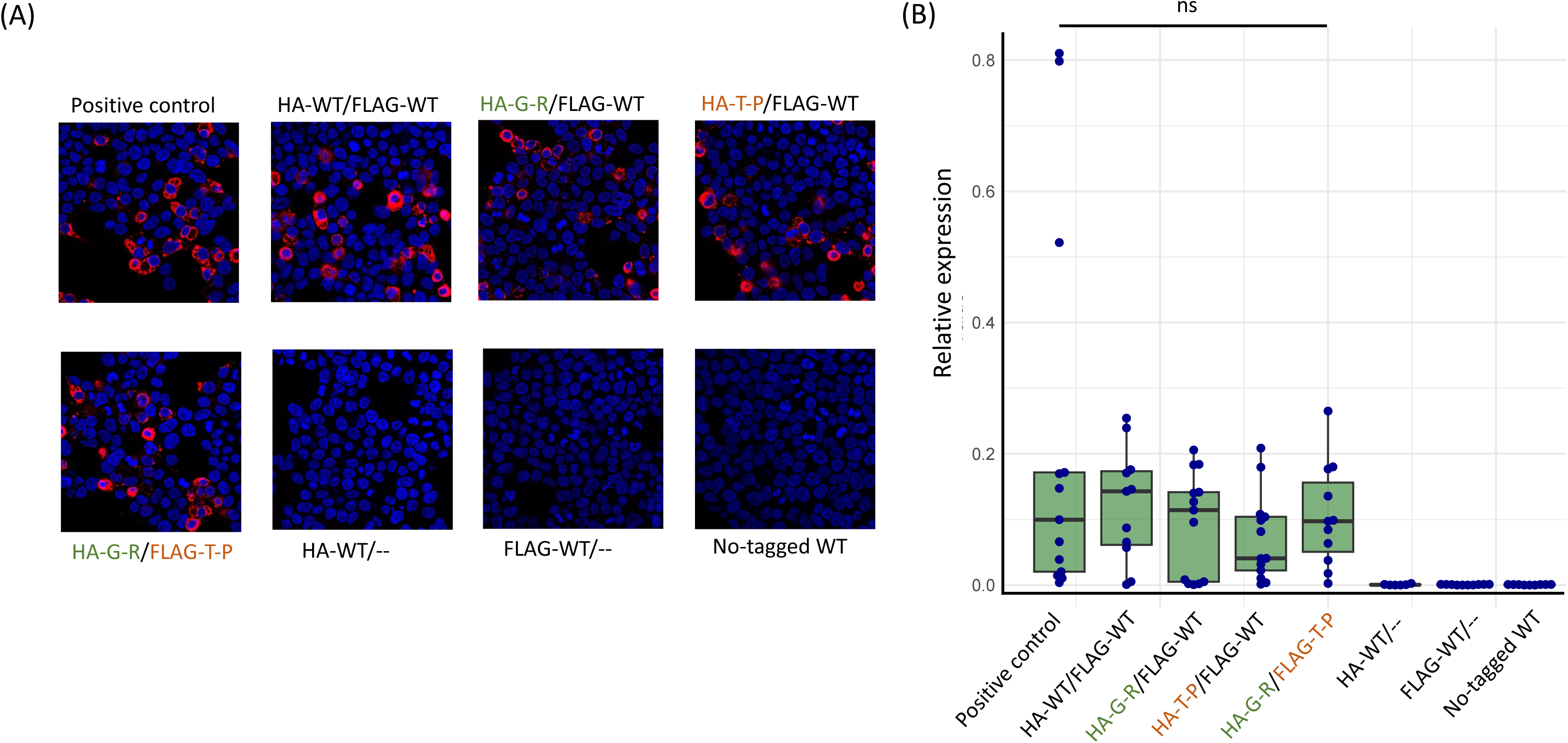
Proximity ligation assay (PLA) A) Microscopic images showing HEK293 cells expressing various plasmid combinations. DAPI was used for nuclei (blue), Duolink proximity ligation signals are shown in red. B) Beeswarm and overlayed box plots representing PLA signal areas normalized by cell numbers (n>9). HA-WT/MT (HA-tagged wild-type/mutant); FLAG-GR/TP (FLAG-tagged G833R/T1396P); HA-GR/TP (HA-tagged G833R/T1396P).

## Discussion

In this study, we identified compound heterozygous *SCN5A* variants in a patient with severe SSS. Functional analysis demonstrated that p.T1396P variant exhibited a severe loss-of-function effect, whereas p.G833R variant retained almost normal function comparable to that of the wild-type (WT) channel. Electrophysiological assays further revealed that the p.T1396P channel exerted a dominant-negative effect on the co-expressed WT channel. Additionally, it delayed current decay and altered the voltage-dependence of steady-state inactivation, suggesting a coupled gating effect.

Notably, all these effects were abolished by co-expression of difopein, consistent with previous reports (ref 6 Clatot 2017). Interestingly, the p.G833R variant was unaffected by the dominant-negative and coupled gating effects. Co-expression of difopein with the variant did not increase current densities.

However, in contrast to this lack of coupling in the p.G833R channel, the co-immunoprecipitation assay confirmed dimerization in all tested channel combinations. In addition, *in situ* proximity ligation assays confirmed the close spatial localization of Na_v_1.5 channels in all tested combinations. These findings indicate that while Na_v_1.5 channels physically interact in close proximity, coupled gating is functionally independent of physical dimerization.

### Distinct roles of Physical Interaction and Functional Coupling in SCN5A

The dominant-negative effect and electrophysiological coupling of loss-of-function *SCN5A* variants have been previously reported *in vitro* and *in vivo* studies^5,6,7^. The hypothesis of Na_v_1.5 dimerization and coupled gating was originally proposed based on electrophysiological findings and co-immunoprecipitation experiments demonstrating physical interaction of two Na_v_1.5 components^5,6^. However, recent evidence suggests that physical dimerization and electrophysiological coupling are distinct. The coupled gating is mediated by 14-3-3 peptide and can be disrupted by difopein, wheareas the dimerization remains unaffected^11^. Our findings align with this, as we observed physical interactions between Na_v_1.5 channels in various combinations, including an electrophysiologically uncoupled variant.

### Clinical insights

Clinically, the degree of loss of function (LOF) of Na_v_1.5 has been associated with disease severity for BrS^12^. However, among these LOF variants, the correlation between the dominant-negative effect and disease severity has yet to be established. Although a previous study reported high penetrance in BrS families carrying DN variants, a clear association with phenotypic severity has not yet been observed^8^. Furthermore, in our analysis of the association between LOF-type *SCN5A* mutations and lethal arrhythmic events (LAEs) in BrS patients, the event rate did not differ between patients with complete LOF missense variants and those with non-missense LOF variants, such as truncation or nonsense variants that likely lack dominant-negative (DN) effects due to haploinsufficiency. Considering that many missense LOF variants have been reported to exhibit DN effects (Oneil et al.), these findings suggest that the presence of a DN effect does not necessarily correlate with worse clinical outcomes^13^.

With a different perspective from these two studies that mainly compare disease severity between carriers with and without DN variants, this study focused on a single family carrying two distinct variants in compound heterozygous fashion. Experimental models that attempt to characterize the phenotype of a single variant both in the presence and absence of DN or coupling effects are not feasible in practice and remain purely theoretical. However, the family in this study represents a rare example in which such a comparison is naturally possible while the phenotype of this family was not BrS rather SSS.

The combination of p.G833R variant, which does not have coupled gating nor DN effects, and the LOF-type p.T1396P variant did not result in a mild phenotype. In terms of current density, p.G833R would be expected to generate a higher density of I_Na_ in the proband’s cardiac tissue, unaffected by the DN effects by p.T1396P. However, in reality, the proband harboring both variants exhibited earlier onset and more severe bradycardia compared to the parent, who carried only p.T1396P. This finding can be explained by two possibilities.

One possibility is the shift of inactivation by p.T1396P. The co-expression of WT and p.T1396P demonstrated a rightward shift of the steady-state inactivation curve via the coupled gating effect. This shift can induce an increase in window current, possibly counterbalancing the reduction of the peak I_Na_ density by the DN effect. In contrast, in the cells expressing p.G833R and p.T1396P, this shift was not observed, suggesting that the proband could not benefit from this compensatory mechanism, resulting in a severe and early-onset phenotype. Another possibility is that coupled gating may occur in normal channels and could play a significant physiological role *in vivo*. Indeed, in our co-expression assay with difopein, not only the cells expressing WT and p.T1396P, but also the cells expressing WT channel alone exhibited a leftward shift in steady-state inactivation, suggesting that WT Na_v_1.5 functions with coupling and that its voltage dependence is naturally rightward-shifted compared to that of the monomer state under normal physiological conditions. In heterozygous carriers of LOF variants and WT allele, the reduced I_Na_ caused by the LOF variant may be compensated by fully functional WT channels. In contrast, in compound heterozygotes with both a LOF variant and a variant lacking coupled gating function, such as p.G833R, the variant lacking coupled gating function may fail to adequately substitute for the LOF channels, thereby exacerbating the disease phenotype. If coupled gating is not a negative factor that causes DN effects, but rather a necessary mechanism for normal channels, then the severe phenotype observed in the proband in this study becomes more understandable.

In any cases, further investigation under physiological conditions is required to elucidate the true impact of the coupled gating effect.

### Research limitations

The electrophysiological and immunological studies were examined only in cultured HEK-293 cells, thus the actual functional characteristics can be different since various channel modulators such as β-subunit, cytoskeleton and membrane proteins exist in human cardiomyocytes. In addition, the genotype of the proband’s sibling was not confirmed.

## Data Availability

The original data supporting the findings of this study are not deposited in a public cloud or repository. However, they will be made available by the corresponding author upon reasonable request.

## Acknowledgement

We would like to thank Ms. Kazu Toyooka, Ms. Miho Goto, Ms. Arisa Ikeda, and Ms. Madoka Tanimoto for their support throughout the study. This study was supported by JSPS KAKENHI 18K15887, 23K07528, a research grant from Mochida memorial fund, Suzuken fund, and Yamauchi-Susumu research fund for KK. During preparation of the work, the authors used Grammarly software and Chat-GPT for English proofreading.

---

**Table.**
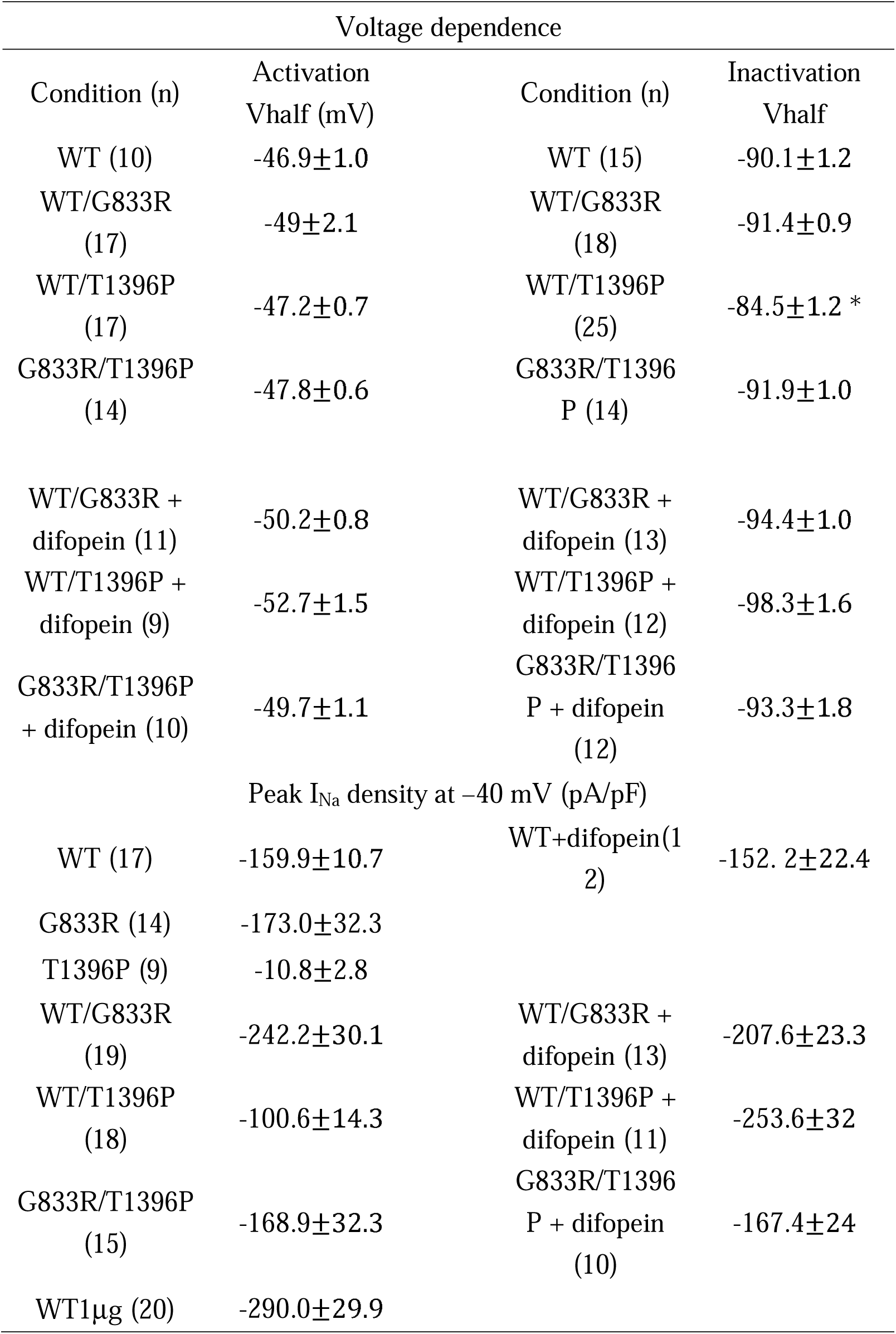
Electrophysiological properties.

## Supplemental Figure

**Supplemental figure legend.**
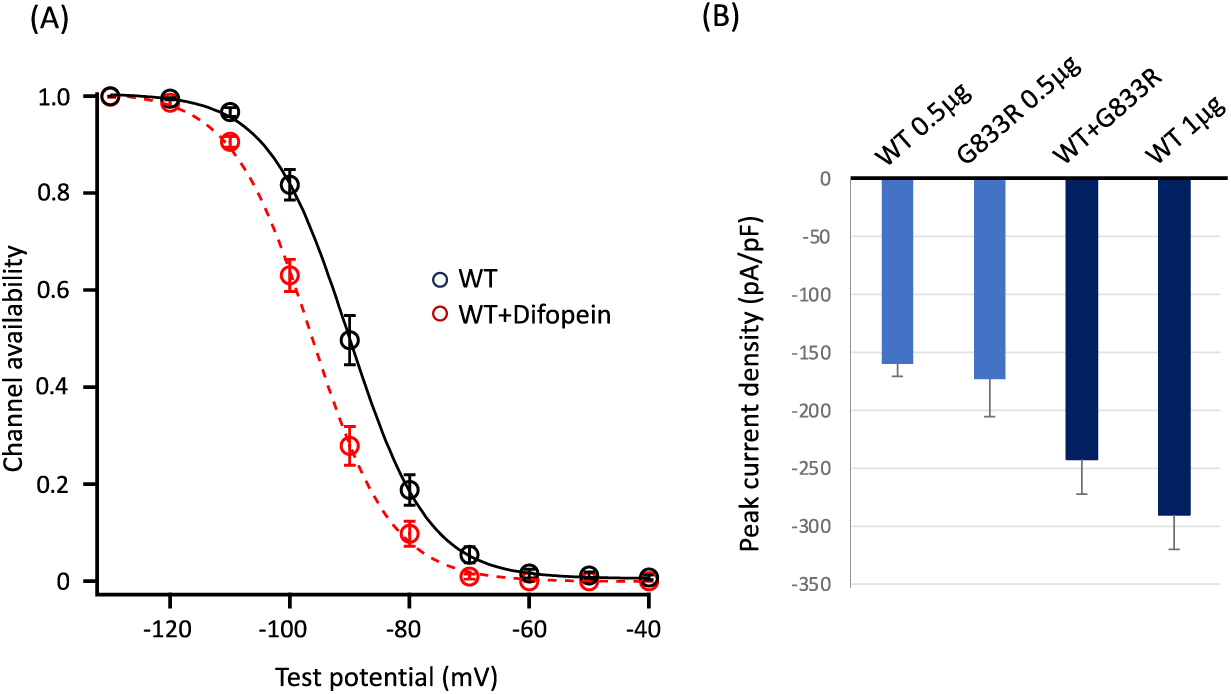
A) Steady-state inactivation curves for cells expressing either WT or WT + difopein. B) Bar graphs representing Peak I_Na_ density at -40mV. Light blue bars indicate cells transfected with 0.5 µg total SCN5A plasmids, while navy bars indicate cells transfected with 1.0 µg total SCN5A plasmids.

